# Development and automated deployment of a specialised machine learning schema within a collaborative research centre: an explorative approach using large language models

**DOI:** 10.1101/2025.10.06.25337418

**Authors:** Klaus Kaier, Gita Benadi, Sophia Nolde, Cristóbal Tagle Ludwig, Claudia Giuliani, Felix Engel, Manuel Watter, Harald Binder

## Abstract

Achieving interoperability in machine learning (ML) workflows remains a significant challenge due to the heterogeneity of data types, algorithms, and application domains, as well as the lack of standardized metadata. In this study, we present the development of a specialized ML metadata schema within the context of the Small Data Initiative, a Collaborative Research Center characterized by diverse scientific approaches. We employed an interdisciplinary process combining expert input, iterative refinement, and schema validation using large language models (LLMs). A two-step LLM-based annotation methodology was applied to 14 representative scientific publications, using six different LLMs to identify both predefined (step 1) and additional ML-related metadata elements (step 2). Manual validation through face-to-face interviews with the main authors to the publications confirmed high precision rates of 70%–85% in the initial step and 86%–98% in the second step, with notable performance variation across models. This approach enabled both the identification of schema inconsistencies and the integration of previously overlooked concepts, leading to the refinement of the metadata schema. The process supports an “AI-by-design” paradigm, ensuring that metadata schemas and annotation workflows are optimized from the outset for downstream AI/ML applications. Our findings also highlight the value of LLM benchmarking in selecting suitable models for domain-specific tasks. Overall, the proposed methodology enhances metadata quality, fosters reproducibility, and contributes to making research data more AI-ready.

## Introduction

Despite efforts in workflow systems and metadata repositories, achieving high interoperability in machine learning (ML) experiments remains challenging [1–3]. This is largely due to different ML platforms having their own specific data and metadata conceptualizations [1, 2]. Furthermore, while initiatives exist, a comprehensive top-level ontology for the entire Machine Learning domain is lacking, with most ontologies focusing on very specific sub-domains [2, 4, 5]. Finally, even with existing ontologies, automatically extracting appropriate metadata from repositories is difficult since most relevant information is not directly available or standardized. Its retrieval thus often requires deduction and domain knowledge [6, 7].

In the present work, we describe the development and automated deployment of a specialised ML metadata schema within the Small Data Initiative (CRC1597), a Collaborative Research Center (CRC) funded by the German Research Association (*Deutsche Forschungsgemeinschaft*, DFG) which is subsequently referred to as SmallData. A major challenge for a joint schema is the diversity of approaches within the CRC: the overall direction of the approach (e.g. clustering, prediction), ML algorithms used (e.g. Bayesian statistics, large language models (LLMs)), and the type of data (e.g. genomics, time-series data). This necessitates a flexible, yet standardized, framework. The metadata schema should accommodate the heterogeneity of SmallData while facilitating knowledge transfer and algorithm comparisons across projects through an automated metadata annotation approach.

In detail, we initiated a multi-step collaborative process led by research data management experts and involving researchers from a representative selection of sub-projects within SmallData: (1) Drawing on existing relevant ontologies and standards [4], an initial draft of the metadata schema was created. (2) This draft was then iteratively refined through feedback loops with participating scientists. (3) Subsequently, the preliminary schema was validated using a LLM-based approach applied to a representative set of relevant scientific publications (N = 14). This last step aimed to identify potentially overlooked ML-concepts or relationships, thereby enriching the schema prior to finalization. (4) To assess the validity of this approach, a supervised double-checking process was conducted using face-to-face interviews.

## Methods

A total of 14 articles published during the first funding year of SmallData were included in the evaluation [8–21]. These articles were selected based on the expectation that they constitute a representative subset of the scientific output relevant to SmallData.

A 2-step approach was used for ML-annotation prediction: In a first step, an LLM was instructed to analyze the full text of an article pdf and only identify ML-annotations that were defined in the preliminary schema. In a second step the LLM was instructed to reanalyze the full text of the manuscript with the task to identify potential ML-annotations that were not covered in step one. See Supplemental Figure 1 for details on the prompting used in the two-step approach. For each manuscript, this two-step approach was repeated for six LLMs. Specifically, we chose to use three so-called ‘reasoning-models’: Perplexity R1 1776 (2025-02-19), Gemini 2.0 Flash Thinking (2025-01-21) and o3 Mini Medium (2025-01-32); as well as three so-called ‘non-reasoning-models’: Gemini 2.0 Pro (2025-02-05), Claude Sonnet 3.7 (2025-02-24) and GPT-4o (2025-01-29). All models were addressed via API calls using OpenRouter. To encourage diverse and exploratory outputs from the language models, we used a temperature of 1, top-p of 1, and top-k of 0 for each LLM.

To assess the accuracy of the predicted ML-annotations in both steps, a supervised annotation validation process was employed. This process involved face-to-face interviews between a research data management expert and a senior scientist who had authored the respective article. Scientists were instructed to evaluate the ML-annotations in the context of the datasets referenced in their publication. Each annotation generated by the two-step approach was reviewed to determine whether it accurately described the ML-approach used in the study. In cases where an annotation was suspected to be inaccurate, participants were encouraged to consult the methods or results sections of the printed article for rapid verification. ML-annotations could also be marked as incorrect based on evident errors or reasonable doubt, without requiring comprehensive cross-checking, in order to maintain efficiency.

To estimate the overall prediction accuracy of the LLMs across all studies, methods for meta-analysis of single proportions were applied [22]. Given the small number of entities per study and the presence of proportions close to 1, the Freeman–Tukey double arcsine transformation was employed [23]. This transformation stabilizes variances and improves the robustness of the meta-analysis under conditions of small sample sizes. A random-effects model using the restricted maximum likelihood (REML) estimator was used to account for between-study variability. The stochastic nature of the LLM, combined with potential variability in how individual authors evaluate the model’s suggestions, further justified the calculation of confidence intervals for the accuracy estimates at the level of the individual articles.

To generate suggestions for possible additions to the metadata schema based on the LLM annotations in step 2 of the approach, we calculated for how many papers and by how many LLMs each item was suggested. In addition, for LLM annotations based on the metadata schema (step 1) we determined the frequency of incorrect suggestion of each item separately for each LLM. In both cases, different spellings, abbreviated and unabbreviated forms and other variants of the same item were standardized prior to the analysis.

SmallData scientists were presented with a list of items not included in the schema that were suggested for at least two papers by all six LLMs combined. The researchers were then asked to decide which of these items should be included in the schema and to identify the appropriate category. In a similar manner, we prepared a list of items that were incorrectly identified in at least two papers by the overall best-performing LLM (o3 Mini). Based on this list and considerations of usefulness for finding relevant ML papers, some items were excluded from the metadata schema.

## Results

The outcomes of the first step are presented in Figure 1a. In this step, the LLMs were tasked with reviewing the full text of a manuscript, focusing exclusively on identifying ML-related annotations as outlined in the preliminary schema. Precision—defined as the proportion of correctly identified entities relative to the total number of annotation suggestions—ranged from 70% to 85%. This indicates that, according to the face-to-face validation interviews, most of the identified items accurately reflected the machine learning approaches employed in the studies. As illustrated in Figure 1a, the highest precision scores were achieved by o3 Mini (85.4%), R1 1776 (83.8%), and Gemini Pro 2.0 (82.7%), whereas Gemini 2.0 Flash Thinking exhibited the lowest precision (71.1%). Details regarding the meta-analysis results on the study level are shown in Supplemental Figure 2.

**Figure 1:**
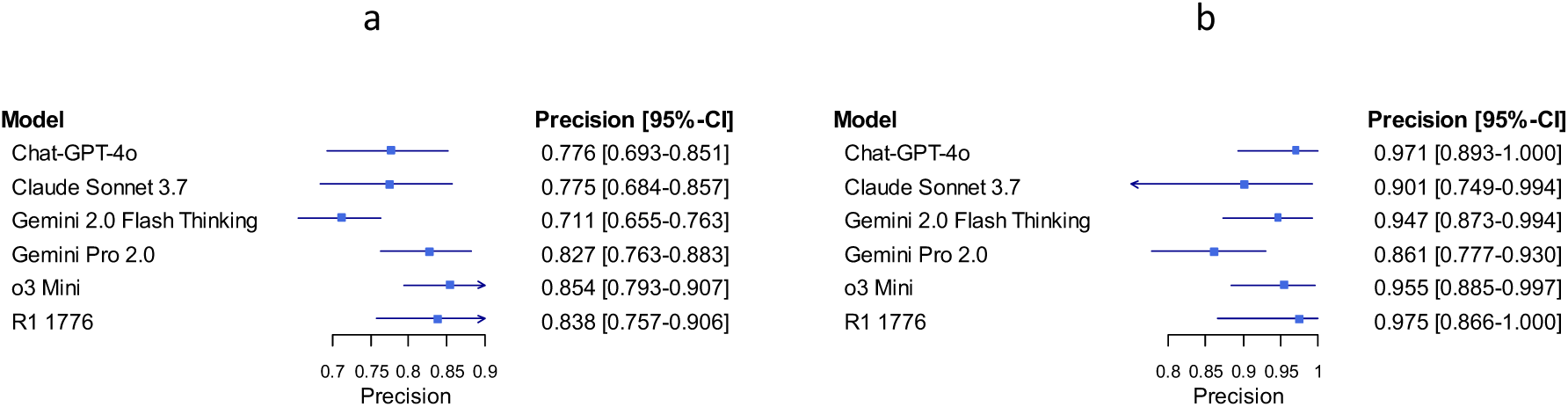
Precision of the different LLMs in step 1 (a) and step 2 (b)

The results of the second step are displayed in Figure 1b. During this phase, the LLMs were tasked with reanalysing the manuscript’s full text to identify additional ML-related annotations that may have been missed in the initial step. Precision scores in this round ranged from 86% to 98%, suggesting that the vast majority of the identified items accurately reflected the machine learning approaches employed in the studies. As shown in Figure 1b, the highest precision was achieved by R1 1776 (97.5%), followed closely by Chat-GPT-4o (97.1%) and o3 Mini (95.5%). In contrast, Gemini Pro 2.0 demonstrated the lowest precision, at 86.1%. See Supplemental Figure 3 for details regarding the meta-analysis results on the study level.

Figure 2 illustrates the Pareto frontier of our methods in terms of precision and the number of correct ML-related annotations. Across both steps, the Gemini models demonstrated the highest level of diligence in proposing ML annotations. In step 1, the average number of correct ML-related annotations was 33.9 for Gemini 2.0 Flash Thinking and 26.4 for Gemini 2.0 Pro. Similarly, in step 2, these models also identified the highest number of additional ML concepts deemed useful, with mean values again reaching 13.0 for Gemini 2.0 Flash Thinking and 13.4 for Gemini 2.0 Pro.

**Figure 2.**
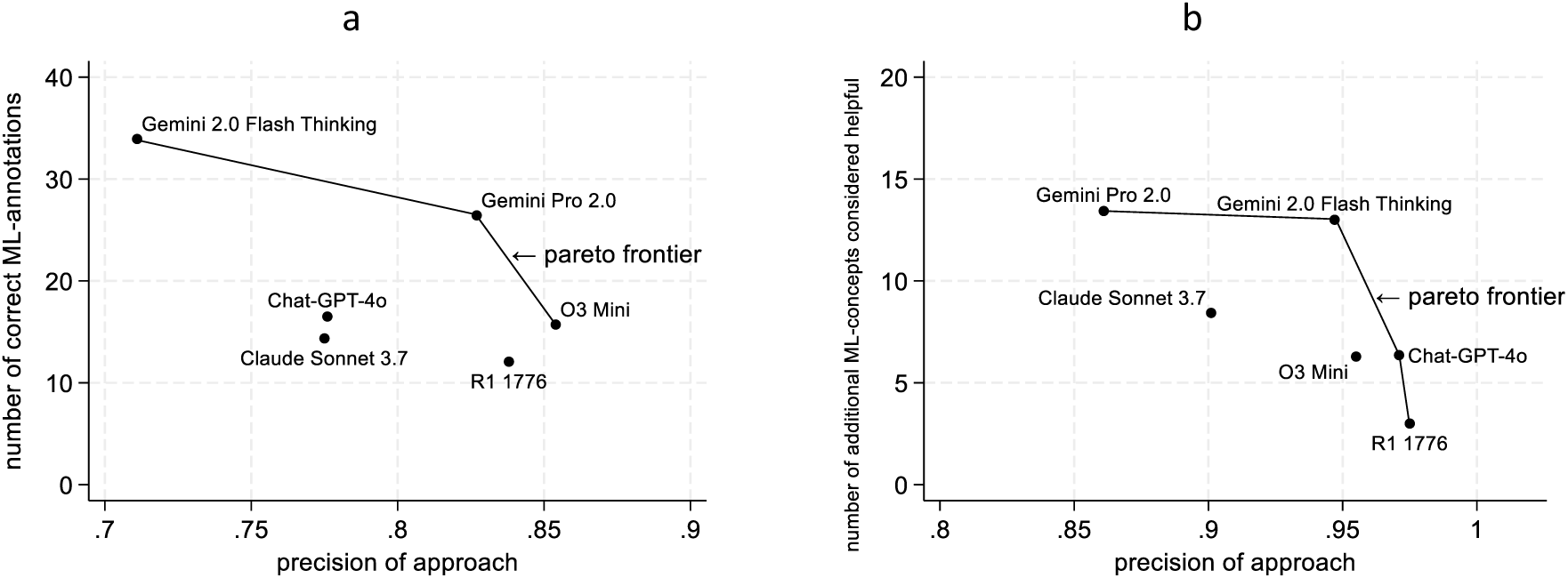
: The Pareto frontier with respect to precision and number of correct predictions in step 1 (a) and step 2 (b) a b

In the detailed analysis of additional aspects of ML identified by the LLMs in step 2 of the approach, 40 items were suggested for two or more papers by at least one LLM (Figure 3, Supplemental Material 4). “Evaluation metrics” was suggested for seven papers by three LLMs, followed by “Loss function” with six papers and two LLMs. In step 1, the LLM with the highest precision, o3 Mini, incorrectly identified nine items in at least two papers (Supplemental Material 5). Four of these items were part of the “Implementation details” category, followed by three items in category “What does it do?”. Based on these analyses, SmallData scientists decided to add seven items to the metadata schema and delete 17 others (Supplemental Material 6). Most deleted items were from the “Implementation details” category.

**Figure 3:**
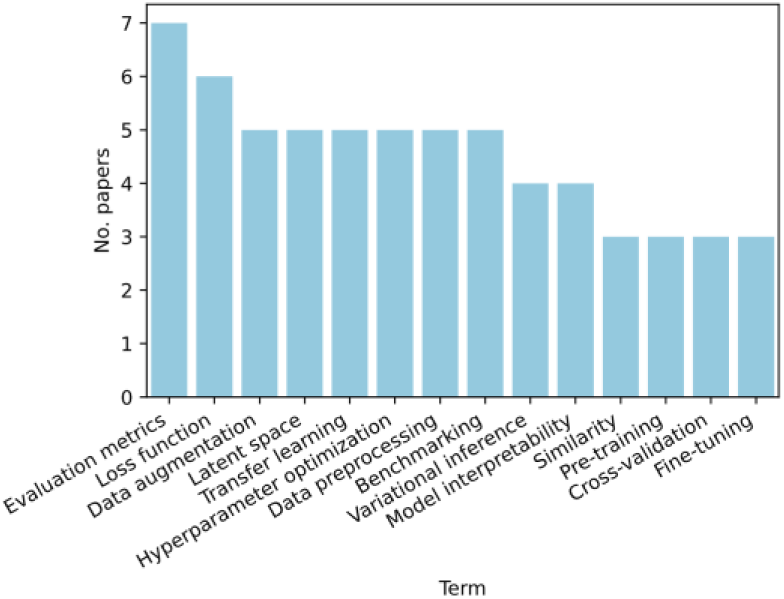
Excerpt of the ML annotations identified in step 2

## Discussion

The development of metadata schemas is now standard in many collaborative initiatives, such as German CRCs, and it is quite common to briefly outline the schema development process when publishing the schema [24]. The LLM-based process we have chosen offers several advantages over a non-LLM-based approach. First, conducting face-to-face interviews with many scientists allows a very diverse insight into what might be relevant. We do not believe that we would have been able to talk in depth about the content of the schema with so many scientists without the pretext of face-to-face interviews. Second, the LLM-based testing of the initial schema (step 1) allows us to identify gross errors in the initial selection of items. Third, the prediction of additional ML annotations (step 2) provides an excellent basis for discussion between research data managers and scientists on what they really consider to be necessary with regard to the annotation of the ML approaches. However, the actual decision as to where the schema should be changed was made in a qualitative manner by the responsible scientists. The role of the research data manager was only to present the quantitatively processed results and to moderate the decision-making pro-cess regarding the changes to the schema. In the end, 17 items were deleted from the initial schema, as they often led to incorrect annotations and/or were not considered appropriate by the scientists responsible. It should be noted that most of the 17 items were removed from the ‘Implementation details’ category. In the preliminary schema, this category lists many aspects of software development that proved to be of little use for the actual use case in the interviews.

An additional advantage of this approach is that it helps make data more “AI-ready” by aligning metadata content and structure with the needs of AI/ML systems. This ensures that metadata schemas and annotation workflows are designed from the outset to support downstream AI/ML use cases—an “AI-by-design” approach.

Last but not least, testing different LLMs gives us important insight into which LLMs are suitable for our specific purposes. As can be seen in Figure 4, there is no visible correlation between the quality criterion used in our specific case (precision) and the most common quality measure for LLMs, the Arena score according to the ranking in the LM Arena [25], imported on 2025-07-06. What’s more, the LLM with the worst Arena score (o3 Mini) actually shows the highest precision measured across both steps.

**Figure 4:**
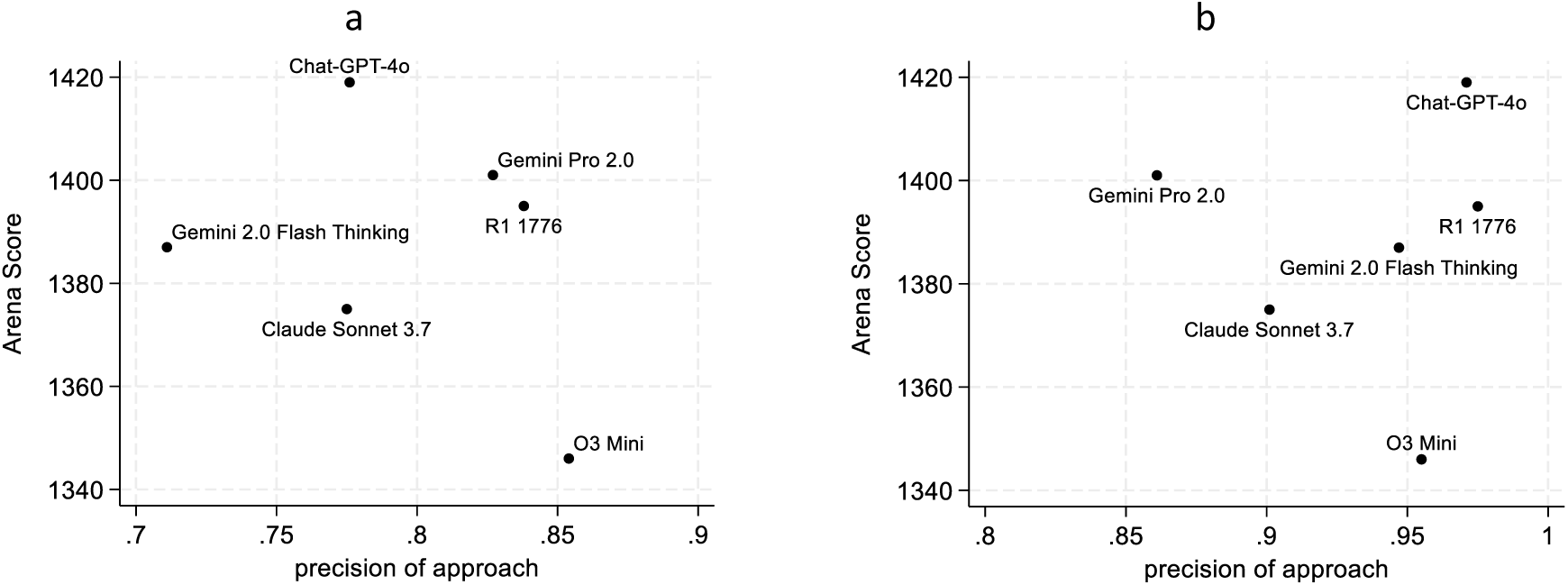
Contrasting the precision of step 1 (a) and step 2 (b) to the Arena Score of the LLMs a b

A major limitation of this approach is its inability to detect ML approaches that the LLMs fail to predict. In other natural language processing studies involving LLMs, false negatives are typically considered when calculating performance metrics such as accuracy, recall, and the F1 score. However, in the present context—where the primary goal is to maximize correct annotations while minimizing false positives—this strategy is not practical [7, 26]. A central challenge is the absence of a clearly defined upper bound for the number of possible annotations in a given article. Another limitation is that the first purpose of the work, to identify the best LLM for future work, conflicts with the second purpose of the work, to further develop the schema based on as many suggestions as possible. On the one hand, we have found that some LLMs are not very accurate when suggesting additional ML annotations. On the other hand, we used the information from all six LLMs to further develop the schema. Here, we must note that in this second step, the suggestions made are only a first opportunity for research data managers to enter into discussions with scientists about further developing the schema. In our case, the final adoption rate is so low that we were grateful for every suggestion. However, this may be substantially different in other contexts. Nevertheless, we note that a variety of LLMs can also be useful, especially in the second step.

Our findings are consistent with other recent efforts that leverage LLMs to automate the extraction of metadata from scientific literature. For instance, a recent study also employed LLMs to populate a predefined schema from research papers, confirming the potential of this technology to streamline metadata creation [27]. However, our approach places a stronger emphasis on a collaborative, “human-in-the-loop” process, using the LLM’s output not as a final product, but as a catalyst for in-depth discussions with domain experts to refine the schema itself. This synergy ensures the resulting schema is not only accurately populated but also conceptually robust and tailored to the specific needs of the research community it serves.

## Data Availability

All data produced in the present study are available upon reasonable request to the authors

---

**Supplemental Figure 1:**
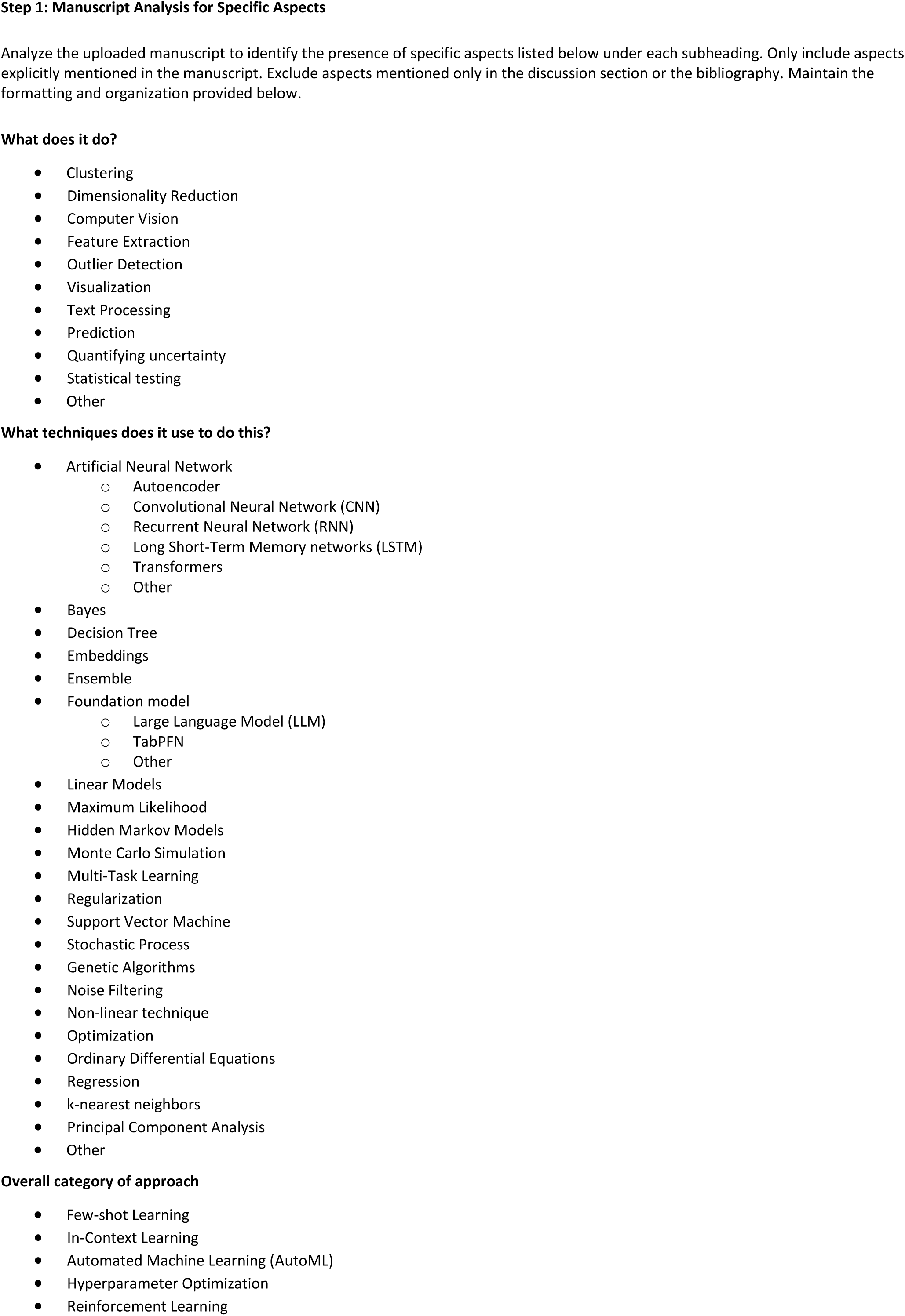
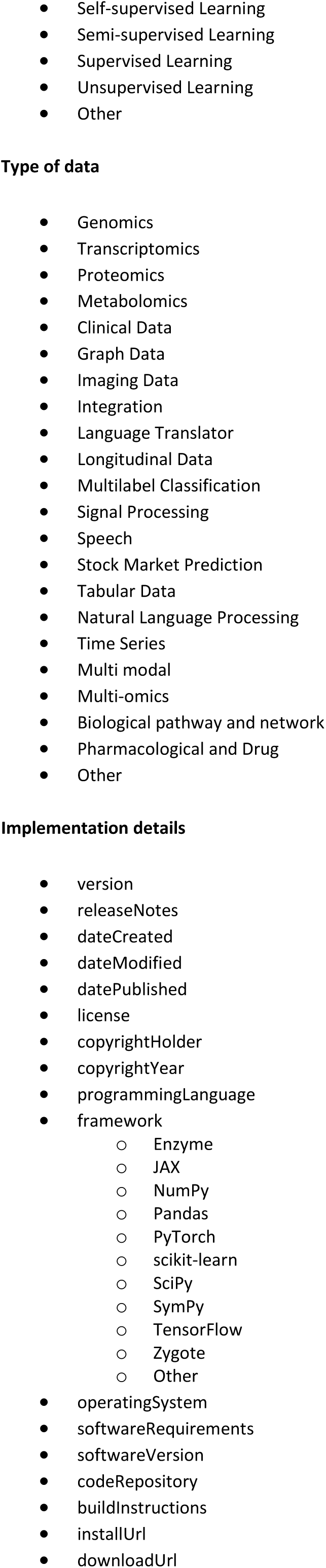
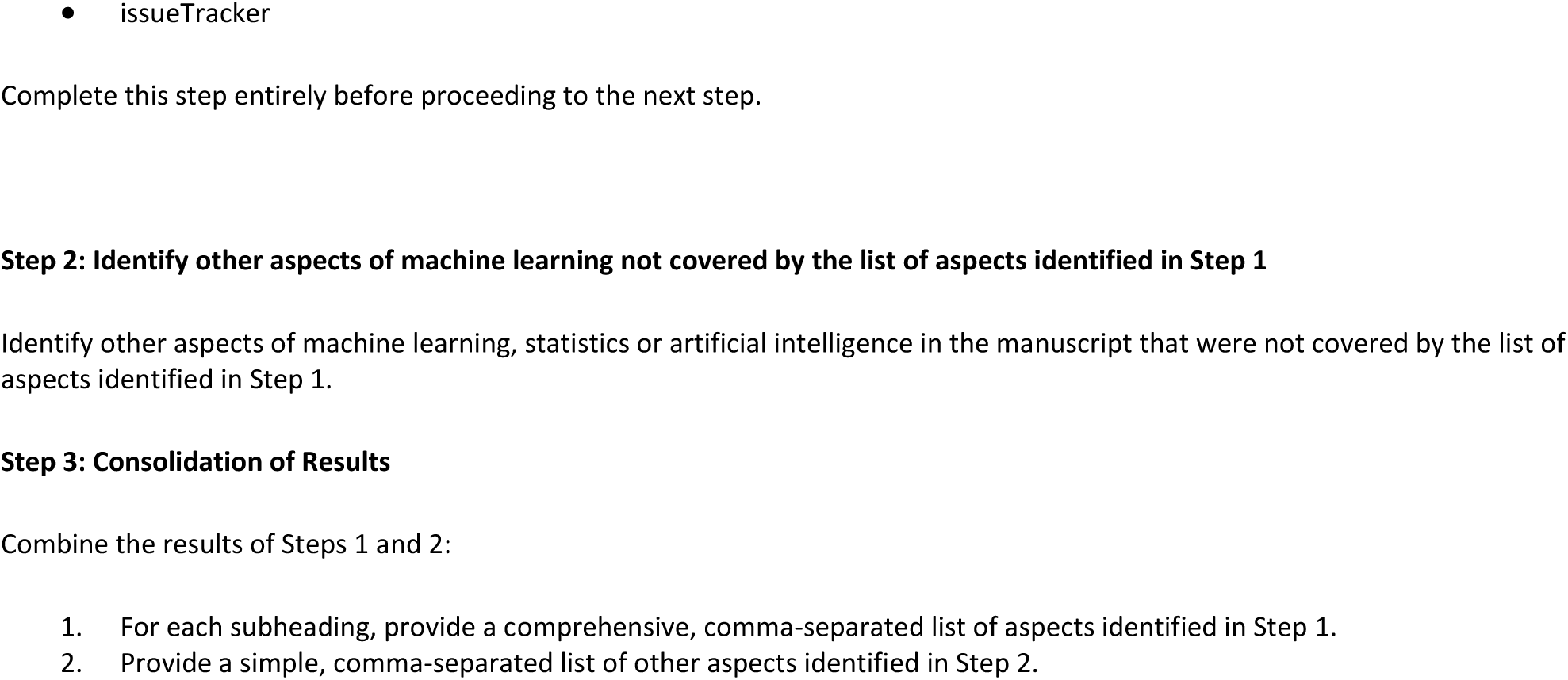
Prompting used in the systematic 2-step approach

**Supplemental Figure 2:**
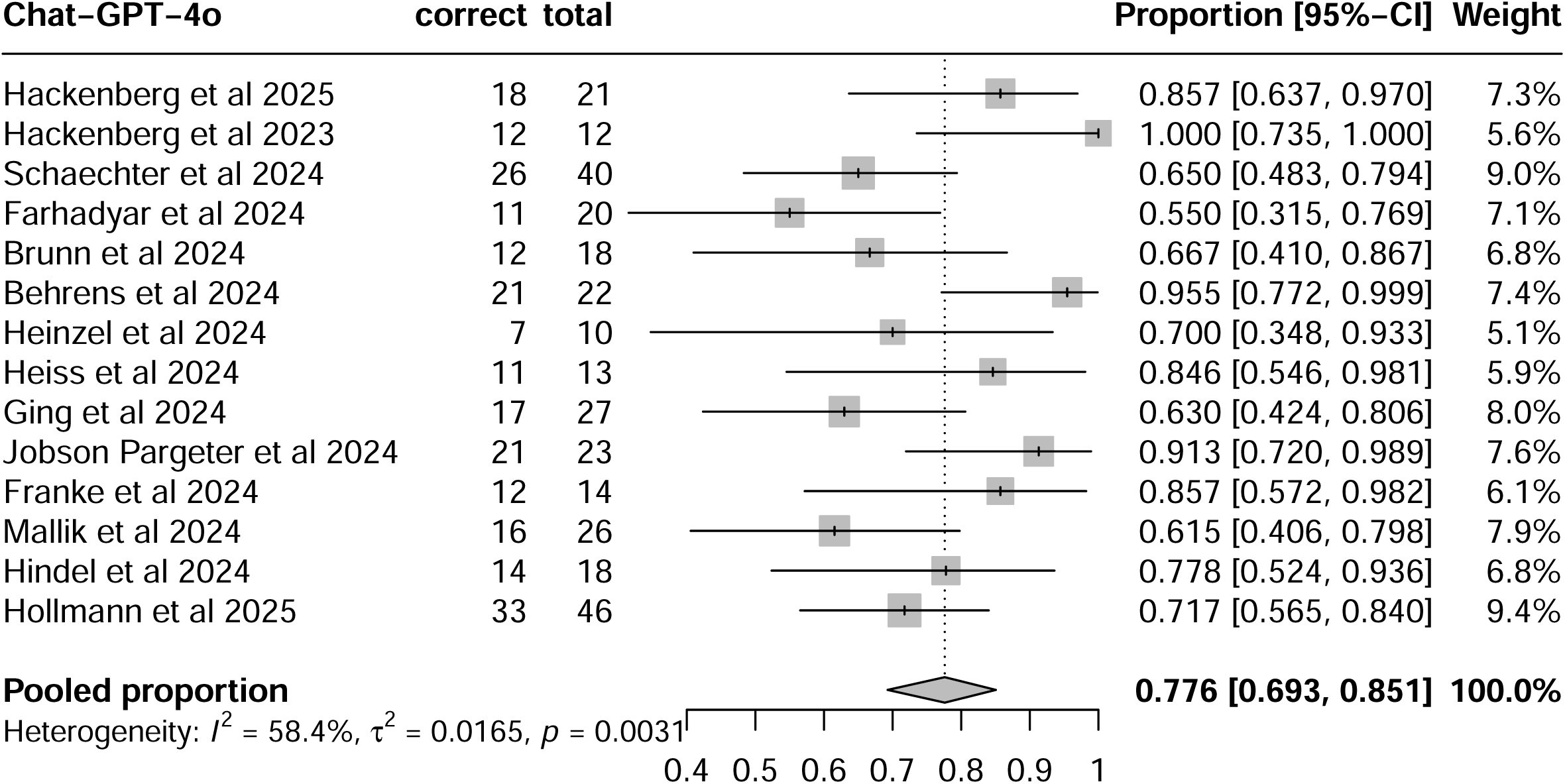
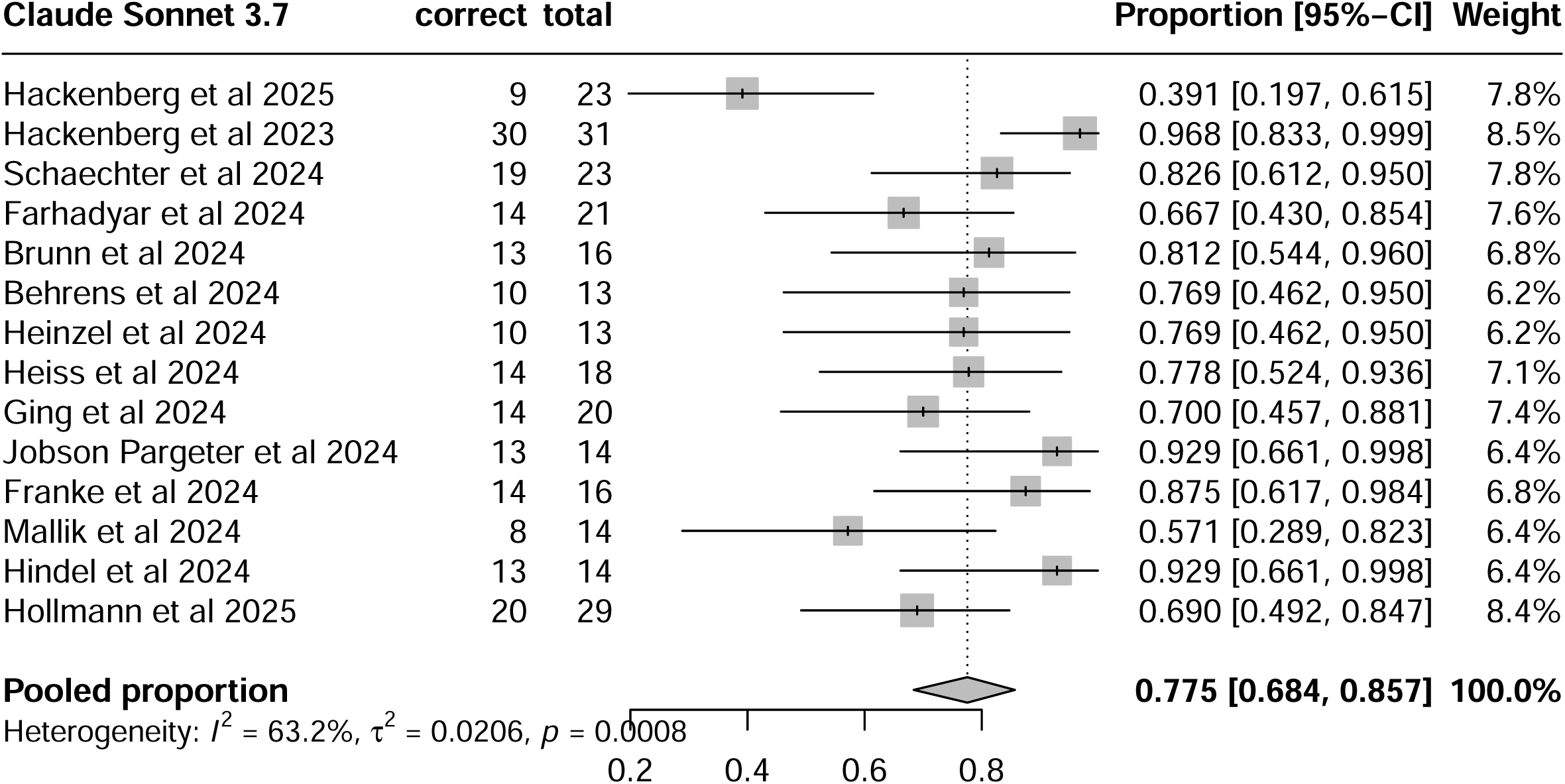
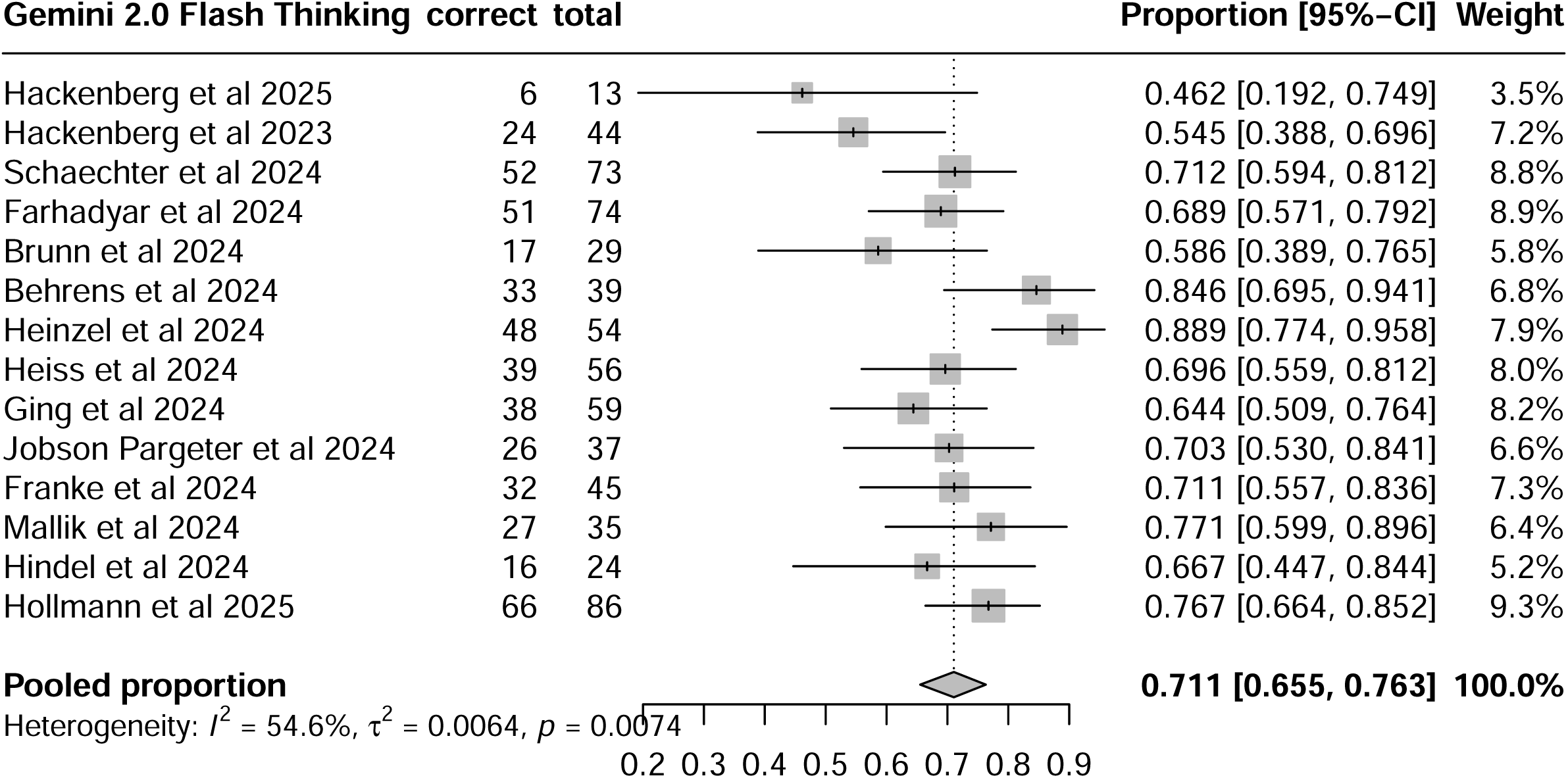
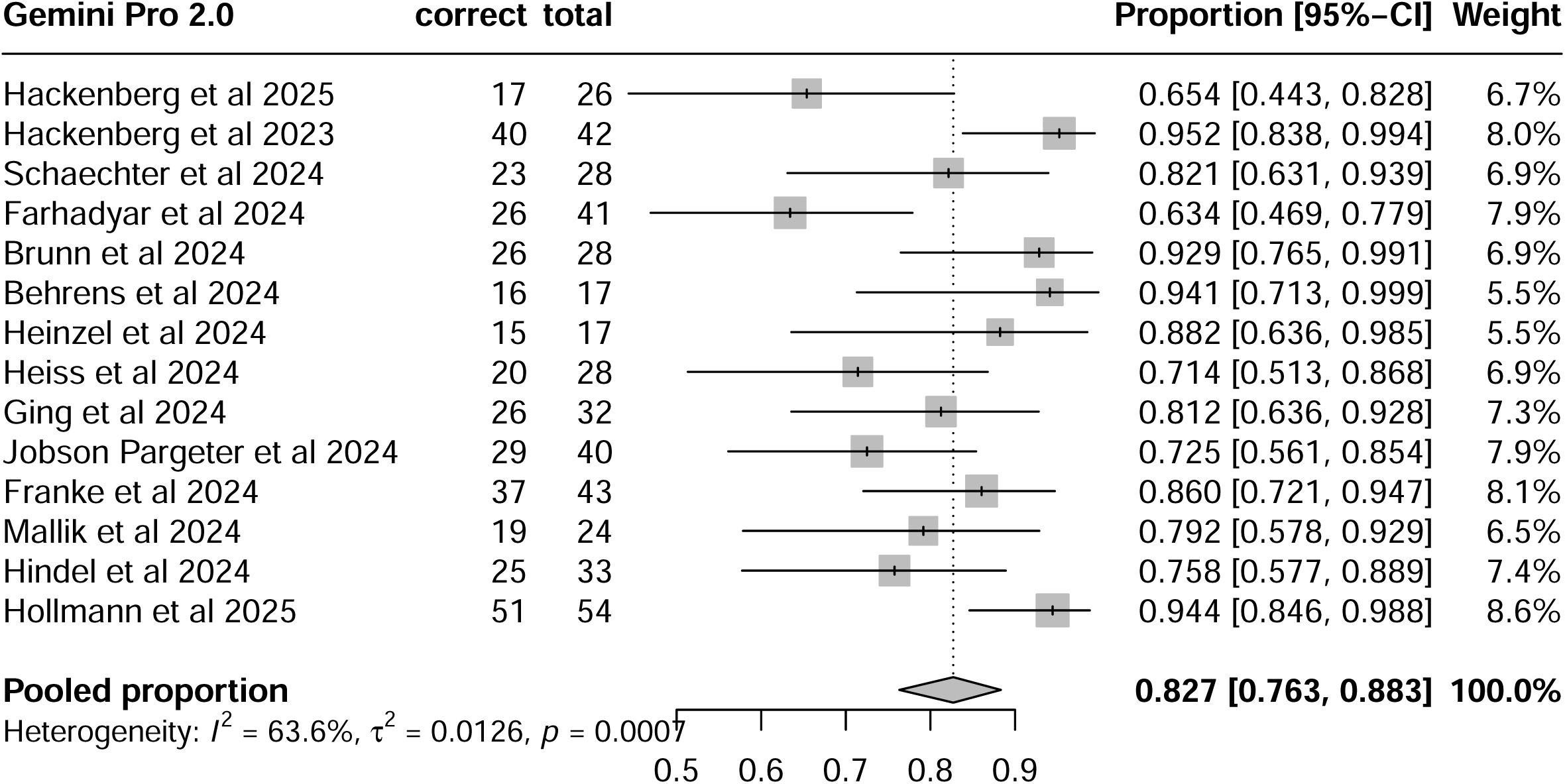
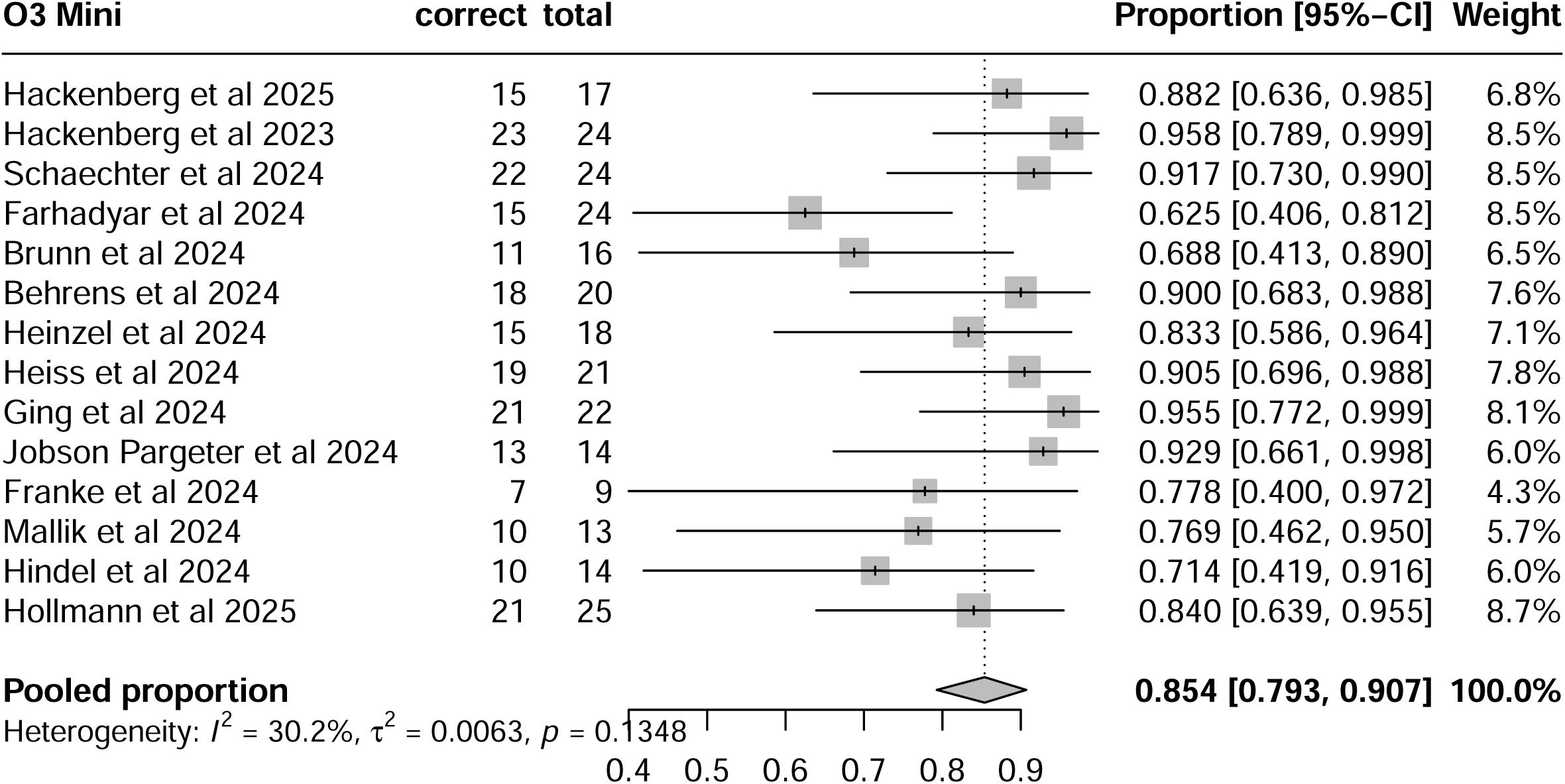
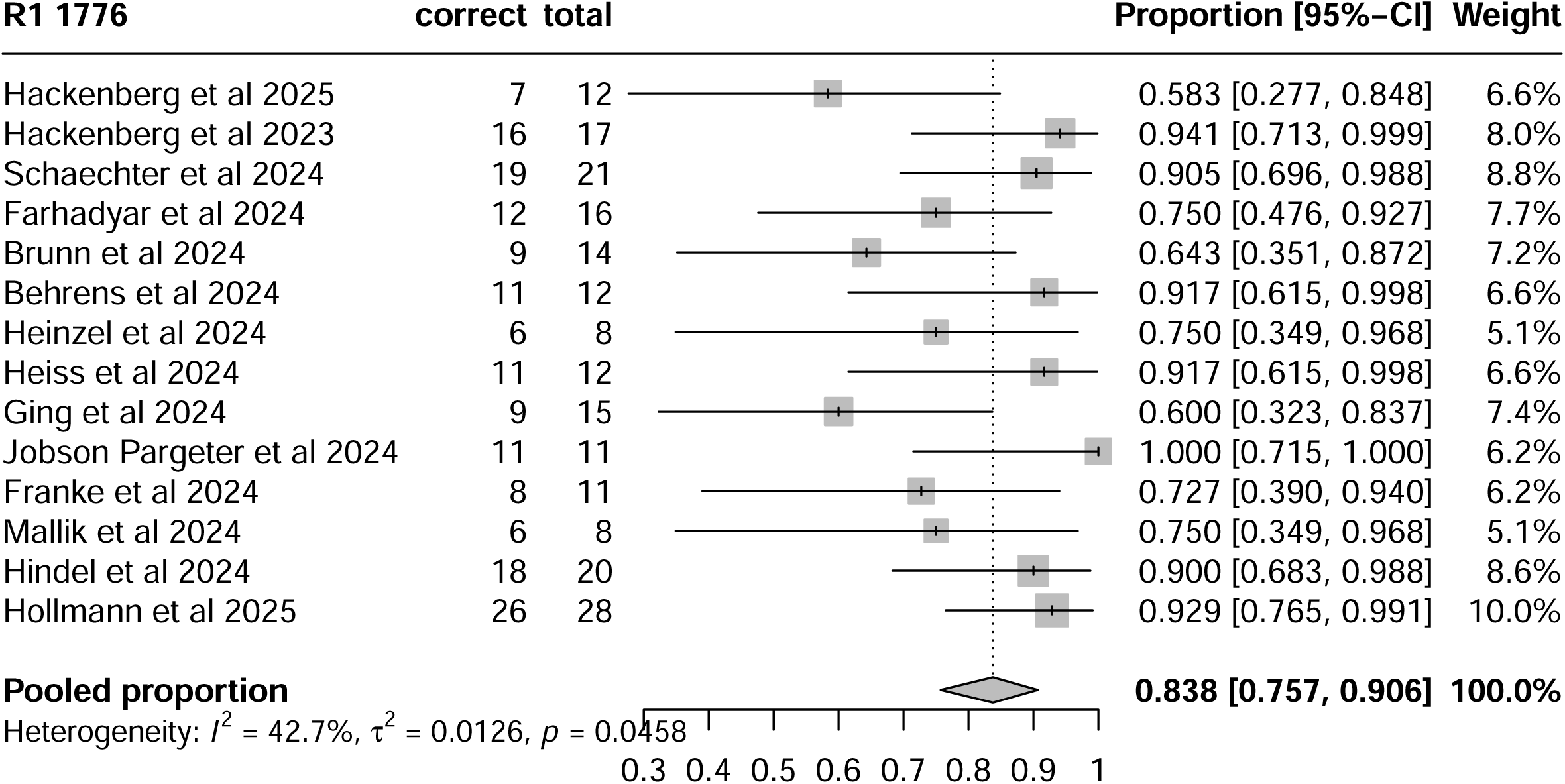
Results of the meta-analyses regarding annotations based on the preliminary schema (step 1)

**Supplemental Figure 3:**
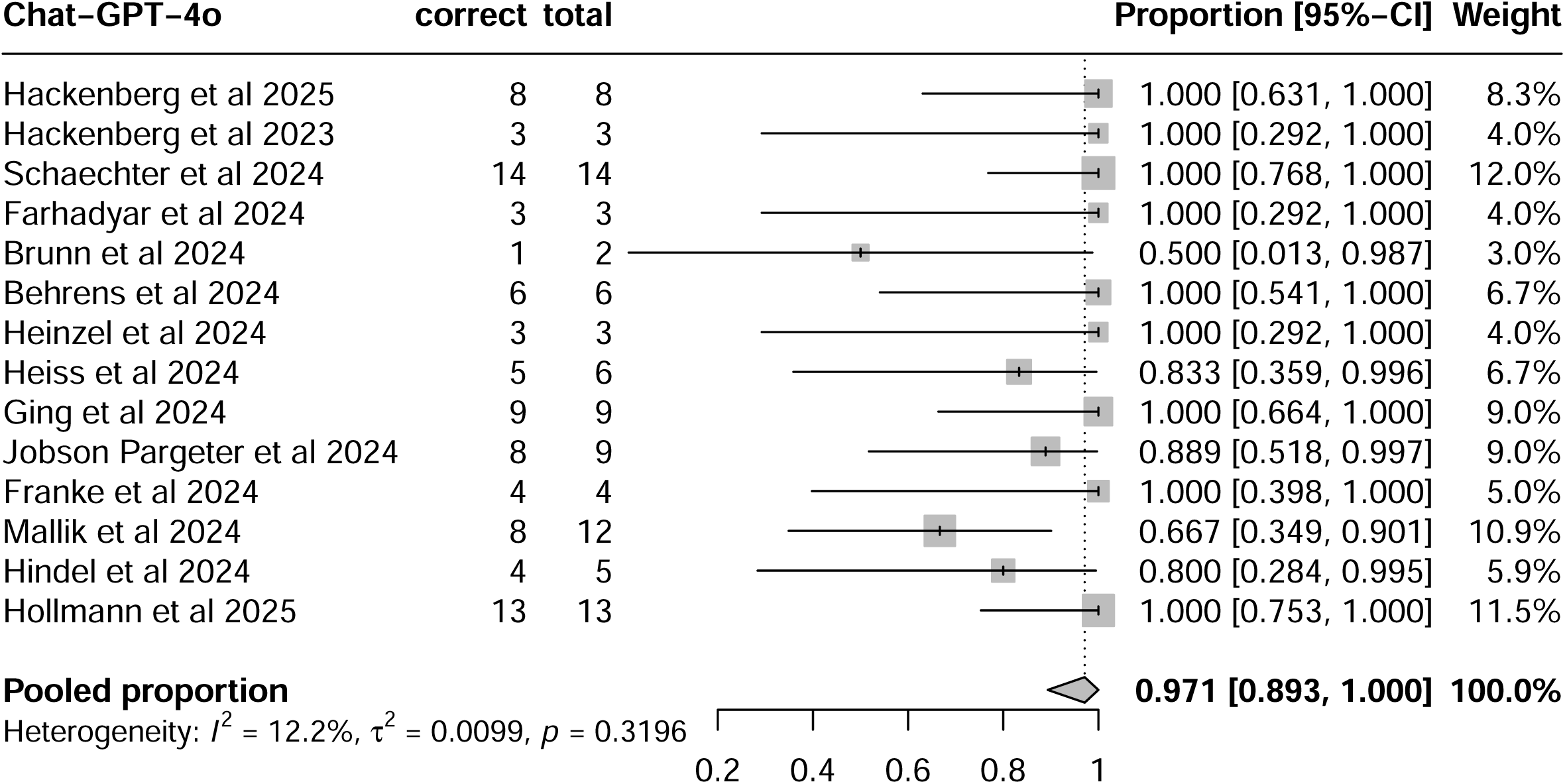
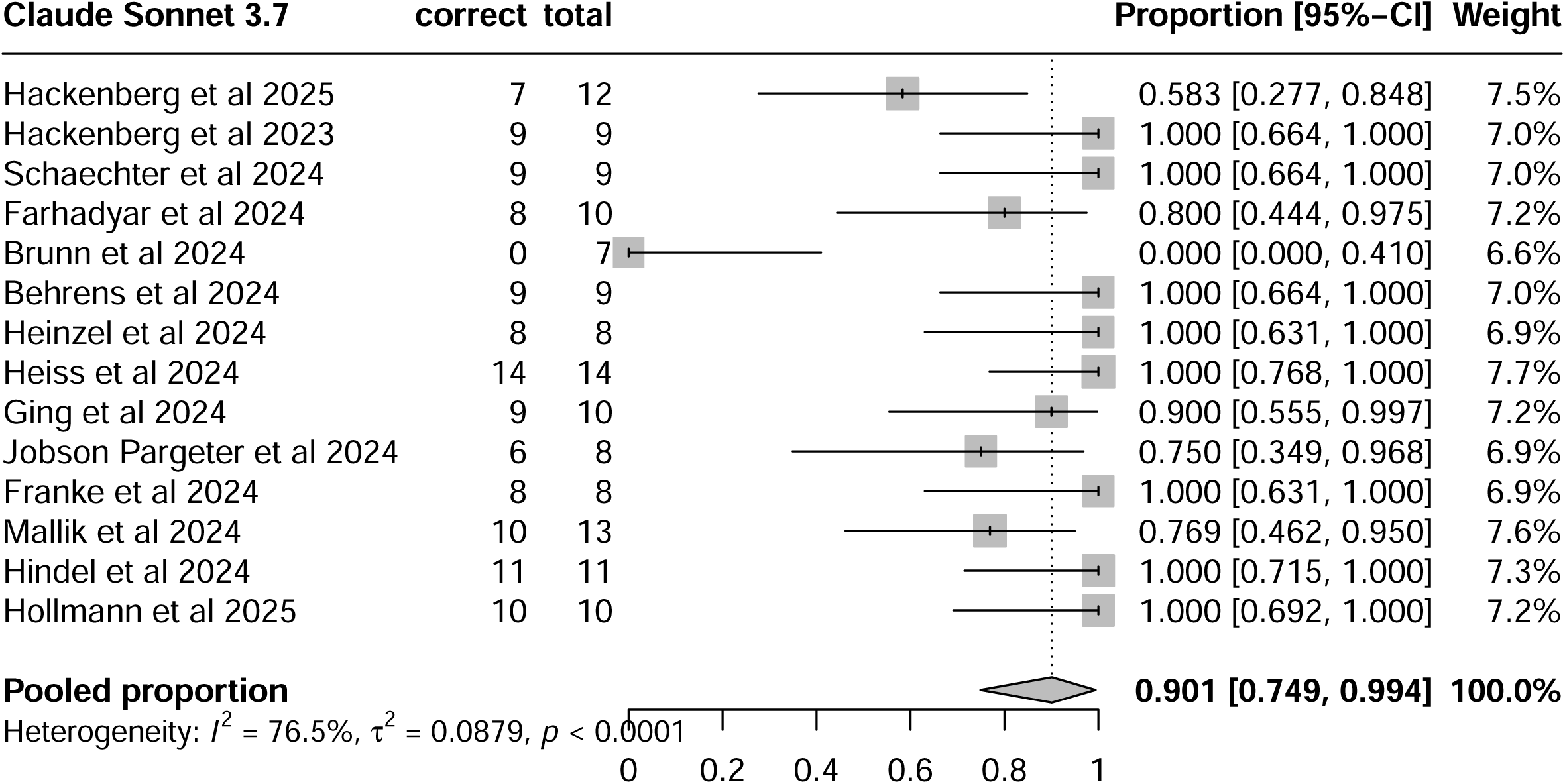
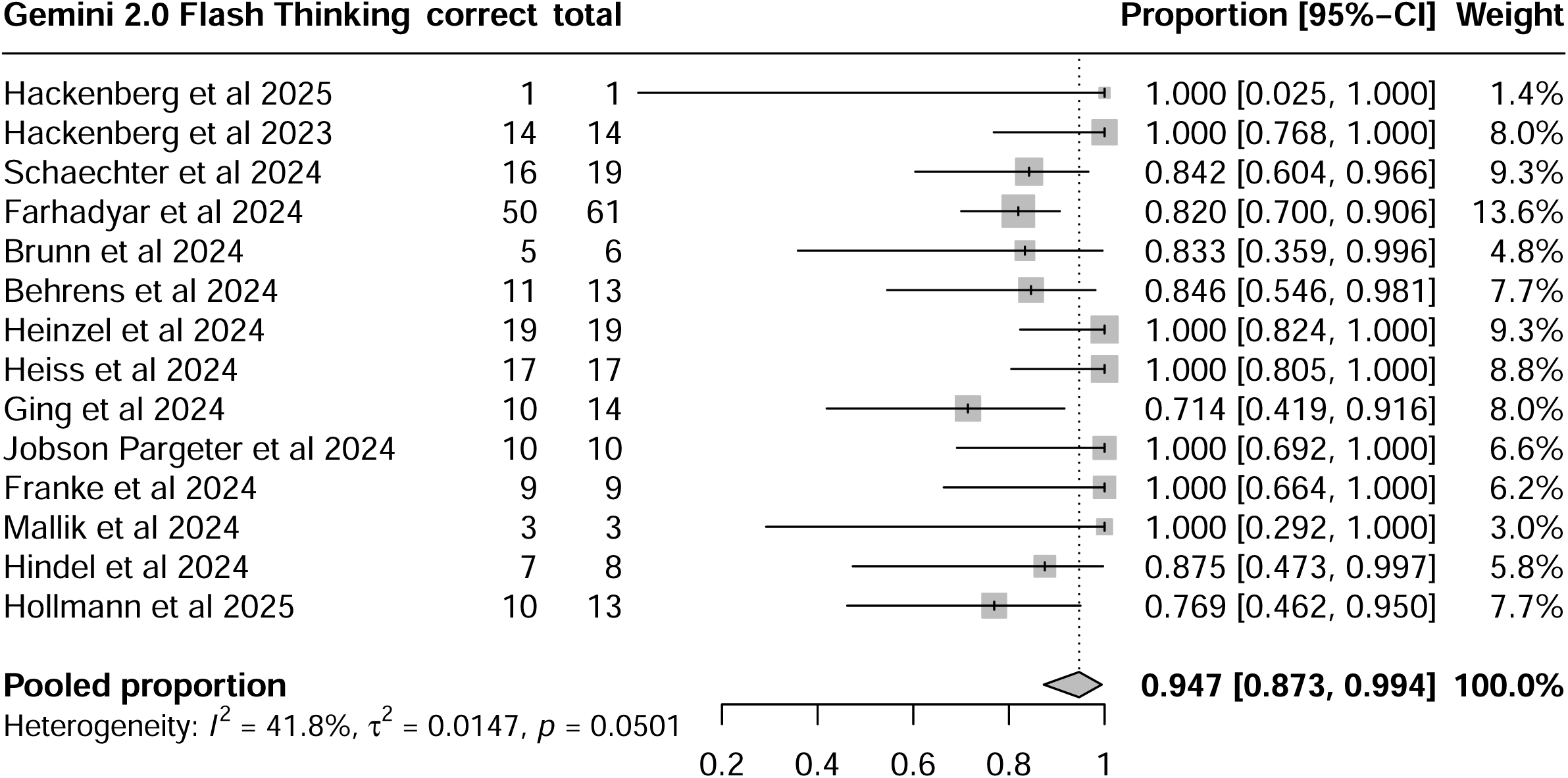
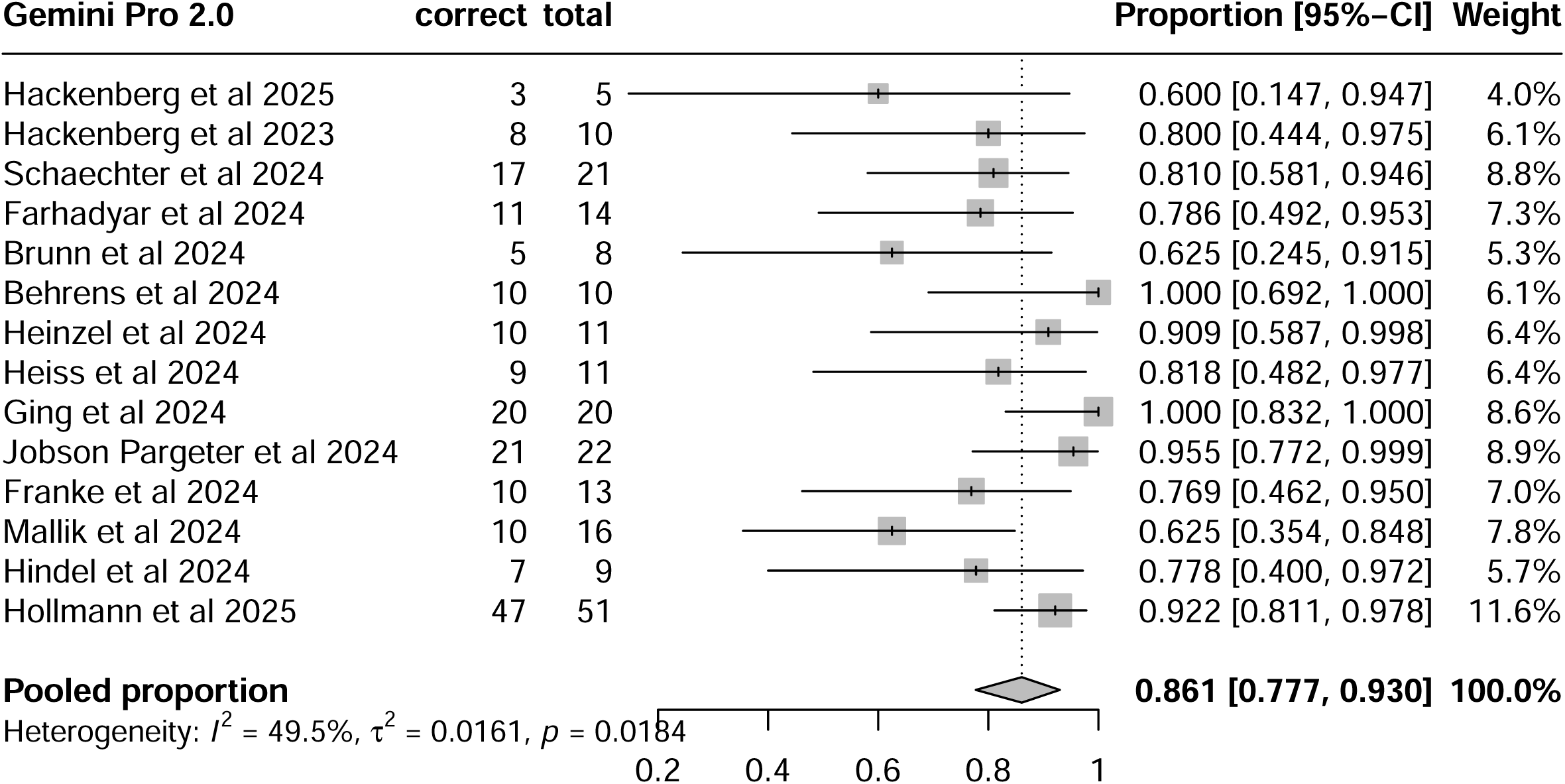
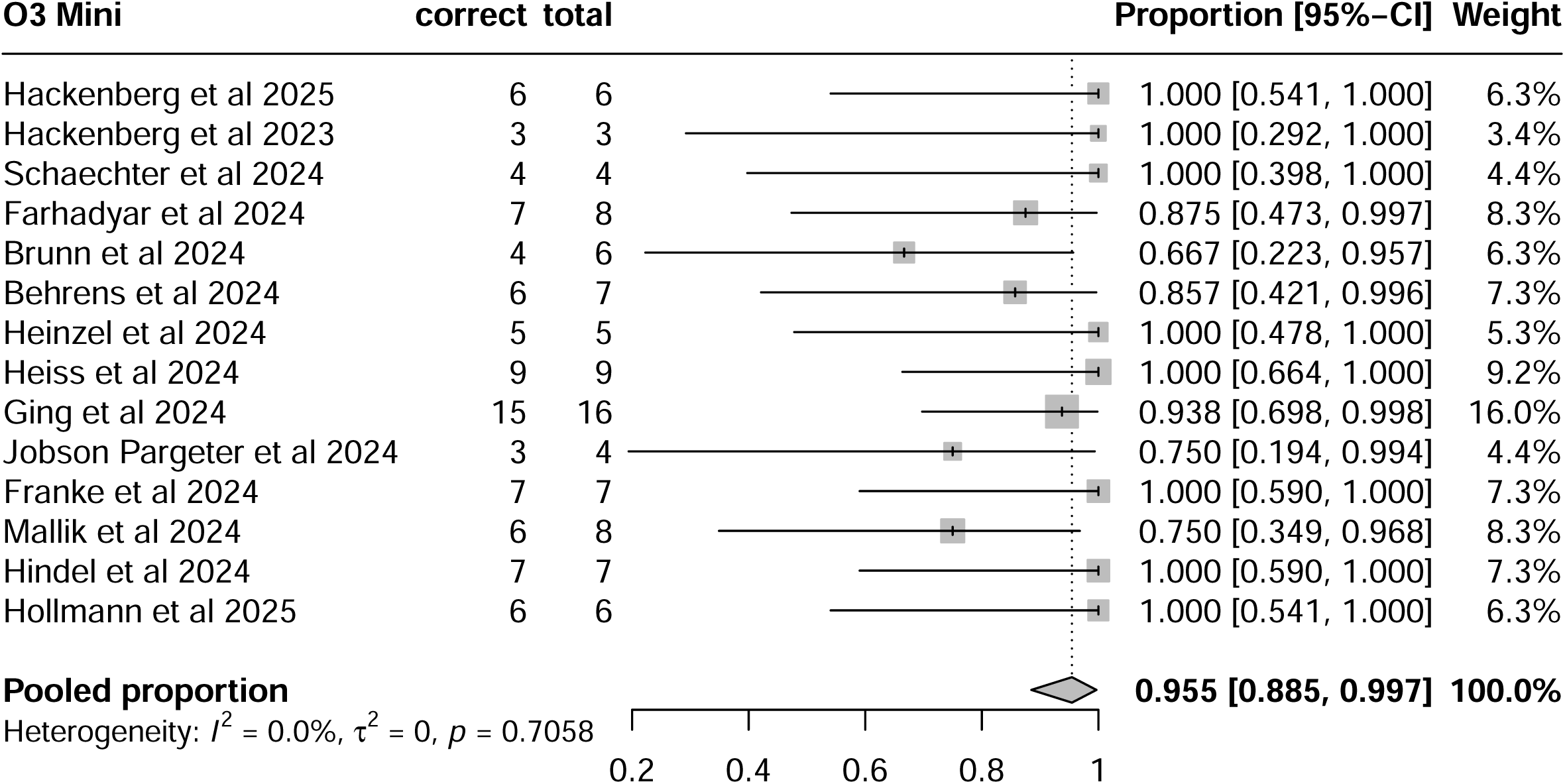
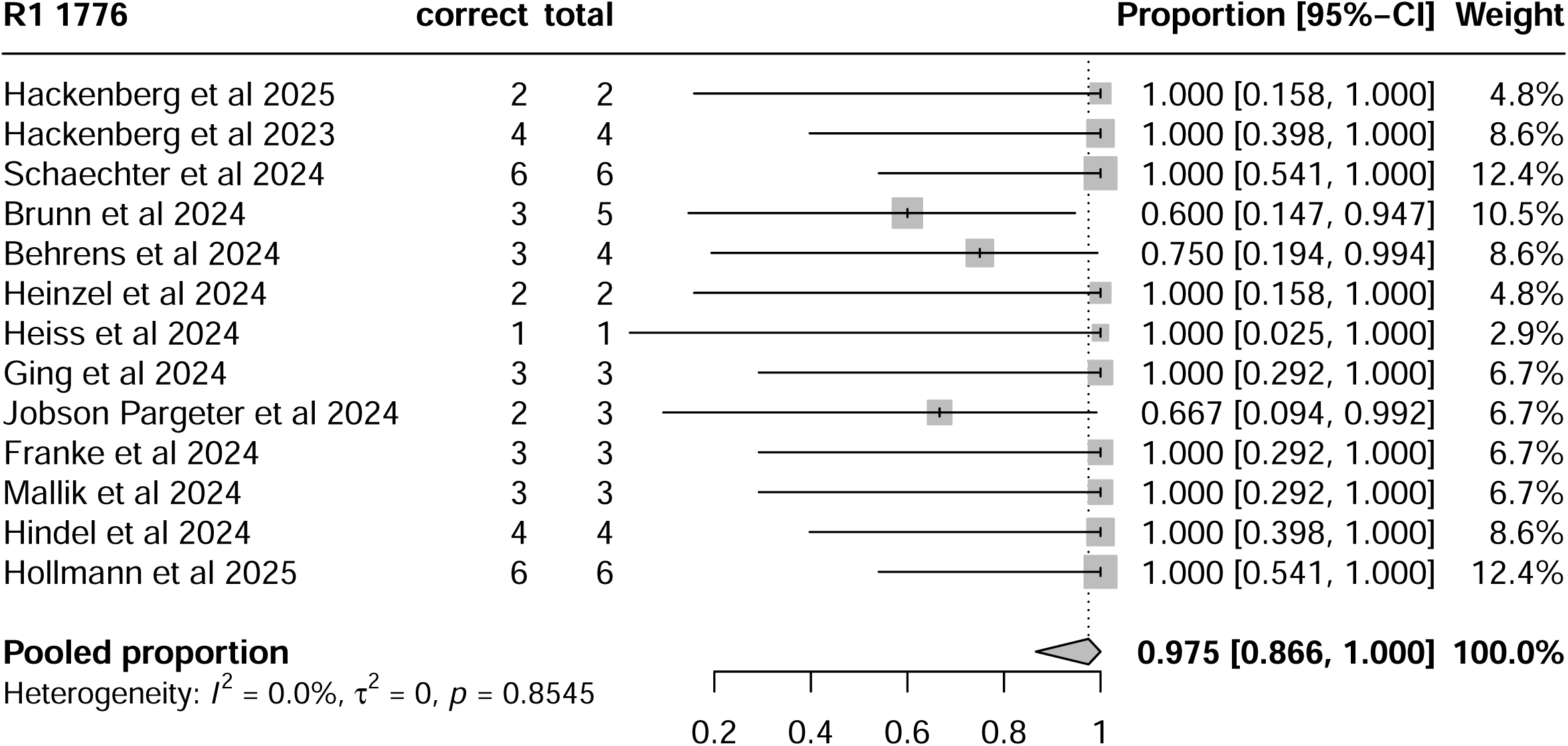
Results of the meta-analyses regarding additional ML-related annotations (step 2)

## Supplemental Material 5

Terms incorrectly suggested by the best-performing LLM (O3 Mini) for at least two papers are marked in **orange**. A number in parentheses indicates the number of papers for which the term was incorrectly suggested.

### What does it do?

- Clustering
- Dimensionality Reduction
- Computer Vision
- Feature Extraction (4)
- Outlier Detection
- Visualization (4)
- Text Processing
- Prediction
- Quantifying uncertainty
- Statistical testing (3)
- Other

### What techniques does it use to do this?

- Artificial Neural Network (3)

- Autoencoder
- Convolutional Neural Network (CNN)
- Recurrent Neural Network (RNN)
- Long Short-Term Memory networks (LSTM)
- Transformers
- Other
- Bayes
- Decision Tree
- Embeddings
- Ensemble
- Foundation model

- Large Language Model (LLM)
- TabPFN
- Other
- Linear Models
- Maximum Likelihood
- Hidden Markov Models
- Monte Carlo Simulation
- Multi-Task Learning
- Regularization
- Support Vector Machine
- Stochastic Process
- Genetic Algorithms
- Noise Filtering
- Non-linear technique
- Optimization
- Ordinary Differential Equations
- Regression
- k-nearest neighbors
- Principal Component Analysis (2)
- Other

### Overall category of approach

- Few-shot Learning
- In-Context Learning
- Automated Machine Learning (AutoML)
- Hyperparameter Optimization
- Reinforcement Learning
- Self-supervised Learning
- Semi-supervised Learning
- Supervised Learning
- Unsupervised Learning
- Other

### Type of data

- Genomics
- Transcriptomics
- Proteomics
- Metabolomics
- Clinical Data
- Graph Data
- Imaging Data
- Integration
- Language Translator
- Longitudinal Data
- Multilabel Classification
- Signal Processing
- Speech
- Stock Market Prediction
- Tabular Data
- Natural Language Processing
- Time Series
- Multi modal
- Multi-omics
- Biological pathway and network
- Pharmacological and Drug
- Other

### Implementation details

- version
- releaseNotes
- dateCreated
- dateModified
- DatePublished (6)
- License (4)
- copyrightHolder
- CopyrightYear (2)
- programmingLanguage
- framework

- Enzyme
- JAX
- NumPy
- Pandas
- PyTorch
- scikit-learn
- SciPy
- SymPy
- TensorFlow
- Zygote
- Other
- peratingSystem
- softwareRequirements
- softwareVersion
- CodeRepository (3)
- buildInstructions
- installUrl
- downloadUrl
- issueTracker

## **Supplemental Material 4:** Changes made in the schema

Terms deleted from the initial schema are marked in red, terms added to the schema in blue.

### 1. What does it do?

- Clustering
- Dimensionality Reduction
- Computer Vision
- Feature Extraction
- Outlier Detection
- Visualization
- Text Processing
- Prediction
- Quantifying uncertainty
- Statistical testing
- Transfer Learning
- Similarity
- Synthetic Data
- Causal Inference
- Other

### 2. What techniques does it use to do this?

- Artificial Neural Network

- Large Language Model (LLM)
- TabPFN
- Other
- Linear Models
- Maximum Likelihood
- Hidden Markov Models
- Monte Carlo Simulation
- Multi-Task Learning
- Regularization
- Support Vector Machine
- Stochastic Process
- Genetic Algorithms
- Noise Filtering
- Non-linear technique
- Optimization
- Ordinary Differential Equations
- Regression
- k-nearest neighbors
- Principal Component Analysis
- Generative Model
- Latent Space
- Domain Adaptation
- Other

### 5. Implementation details

- Version
- releaseNotes
- dateCreated
- dateModified
- DatePublished
- License
- copyrightHolder
- CopyrightYear
- Programming Language
- Framework

- Enzyme
- JAX
- NumPy
- Pandas
- PyTorch
- scikit-learn
- SciPy
- SymPy
- TensorFlow
- Zygote
- Other
- OperatingSystem
- softwareRequirements
- softwareVersion
- Code Repository
- buildInstructions
- installUrl
- downloadUrl
- issueTracker

